# Variation in Racial and Ethnic Representation of Heart Transplant Recipients, Waitlists, and Local Census Demographics Across Transplant Centers in the US

**DOI:** 10.1101/2022.03.15.22272420

**Authors:** Edward Thomas

## Abstract

As the uses and needs for heart transplants around the world continue to rise, it’s vital to investigate how the recent boom in demand comparative to availability affects racial and ethnic groups, especially in the representation in heart transplant waitlists and recipients. Here, comparisons between the racial/ethnic representation in heart transplant waitlists and recipients at heart transplant centers through the Scientific Registry for Transplant Recipients (SRTR) are drawn, comparative to the racial/ethnic representation in the location that these centers are located. Findings point towards the overrepresentation of White individuals in both waitlists and recipients, and underrepresentation in all other ethnic minorities --except African Americans-- in waitlists and recipients comparative to their demographic representation in the location of the centers. This difference in representation is largest for Hispanics/Latinos and those that the SRTR classify as “Other”. Though just a snapshot of representation between 07/01/2019 and 06/30/2020, this may point to various systemic issues in the accessibility of care of minorities, especially during the COVID-19 pandemic, that need to be addressed as the general populous ages and heart transplants are increasingly relied upon as a treatment for cardiovascular conditions and failure.

## Introduction

The heart transplant is a recent advancement in medicine, first successfully performed in humans around 50 years ago. After Dr. Christiaan Nethling Barnard performed the first successful human-human transplant from Denise Darvall to Louis Washkansky, who was in end-stage ischemic cardiomyopathy and lived for 18 days post-operation (Stolf, 2017), heart transplantation began to etch its name into medicine as a way to prolong life like never before. However, after an initial climb, the number of heart transplants per year has remained relatively the same since the mid-1990s (Stehlik et al, 2011) but the need for heart transplants has continued to rise and appears to continue to rise (Tonsho et al, 2014; see Fuchs et al, 2019; Hunt & Haddad, 2008) to such an extent that the number of available, transplantable hearts far outweighs the number needed for patients (Stehlik et al, 2018). At the same time, the survival of heart transplants has drastically improved; in the 1970s, survival rates were around 30% but have risen to nearly 90% by the 2000s (Tonsho et al, 2014).

As heart transplants popularize around the world, it’s crucial to begin investigating how various factors may contribute to the onset, progression, and treatment of cardiovascular conditions. As new literature surrounding racial and ethnic disparities in health and healthcare emerge every day, it’s similarly vital to understand how race and ethnicity may influence the prevalence of heart transplants in different communities and risk factors.

Race and ethnicity have been found to play an important role in healthy living as a whole and successful heart transplants post-operation: Black individuals are more likely to be transplanted in transplant centers with poorer performance (Kilic et al, 2015) and generally have higher mortality and morbidity rates than other races (Williams & Rucker, 2000). Native Americans and Hispanics are at higher risks for disease and death rates for a variety of conditions (Williams & Rucker, 2000) and minorities are also less likely to have access to high-quality healthcare in the US because of historically higher rates of unemployment and lower rates of employment in roles that provide health insurance in their employment packages (Blendon et al, 1989). And, even then, cross-racial awareness of health disparities is low (Benz et al, 2011), which can lead to issues in the allocation, distribution, and use of resources to make healthy living equitable and accessible. Yet, little is known about how race and ethnicity influence the likelihood of an individual to enter the heart transplant waiting list and recieve a transplant.

Here, racial/ethnic representation on transplant recipients, waitlists, and the average racial and ethnic representation across the localities of transplant centers, is compared to reveal potential disparities in access to treatment/healthy living, cultural hesitancy towards treatment, and the degree to which inequities may be across the US.

## Methodology

Using the Scientific Registry of Transplant Recipients, all 146 transplant centers in which patients were placed onto the waitlist for a heart transplant/received a heart transplant from 07/01/2019 to 06/30/2020 were identified. Using the Scientific Registry of Transplant Recipients’s Complete Report for each transplant center, data on the demographic percentages of New Waiting List Registrations and of those who were a Deceased Donor Transplant Recipients for the period were collected. In addition, the location of the transplant center was also collected.

Transplant centers that had 3 or fewer individuals that were placed on the waitlist or who underwent a heart transplant were removed from the analysis to prevent skewing of demographic representation due to few or no waitlists/transplants. There were 130 final centers that passed trimming and were analyzed.

The demographics from the United States Census Bureau QuickFacts “Race and Hispanic Origin” Information for the locations that these 130 transplant centers were located in was also recorded. This demographic data was collected on the days of 07/01/2021 and 07/02/2021; no known updates to the data occurred during data collection.

Data from both the Scientific Registry of Transplant Recipients was averaged across all transplant centers and data the United States Census Bureau QuickFacts “Race and Hispanic Origin” Information was averaged across all locations of the transplant centers that were analyzed to provide a single snapshot-glimpse into the general demographic representation of those across all heart transplant waitlists, those who underwent a heart transplant, and their representation in the census in the locations of the centers.

Multiple ANOVA and tTest analyses were then conducted on these data sets across and within racial groups in relation to waitlist, transplant, and census representation using XLMiner Analysis ToolPak.

Note: The SRTR categorizes ethnicity/race into 6 groups: “White”, “African-American”, “Hispanic/Latino”, “Asian”, “Other”, and “Unknown”. The SRTR categorizes those who are “Hispanic/Latino” as their own category which does not overlap with racial groups. No transplant center reported having waitlisted or transplanted a heart for any individual(s) who were under the “Unknown” category from 07/01/2019 to 06/30/2020. As such, the “Unknown” category was eliminated from the analysis.

The United States Census Bureau QuickFacts “Race and Hispanic Origin” Information categorizes race/ethnicity into 8 groups: “White alone”, “Black or African American alone”, “American Indian and Alaska Native alone”, “Asian Alone”, “Native Hawaiian and Other Pacific Islander alone”, “Two or more races”, “Hispanic or Latino”, and “White alone, not Hispanic or Latino”.

To compare the two sources, those that were “Asian alone” in the United States Census Bureau QuickFacts “Race and Hispanic Origin” Information were considered “Asian”.

Those that were “Hispanic or Latino” in the United States Census Bureau QuickFacts “Race and Hispanic Origin” Information were considered “Hispanic/Latino”. Those that were “Black or African American alone” in the United States Census Bureau QuickFacts “Race and Hispanic Origin” Information were considered to be “African-American”. Those that fell under “American Indian and Alaska Native alone”, “Native Hawaiian and Other Pacific Islander alone”, or “Two or more races” in The United States Census Bureau QuickFacts “Race and Hispanic Origin” Information were grouped under “Other”.

As the category “White alone” in the United States Census Bureau QuickFacts “Race and Hispanic Origin” Information overlaps with those in the “Hispanic/Latino” group, the “White Alone” category was not considered for analysis. Instead, those that were part of the “White alone, not Hispanic or Latino” category in the United States Census Bureau QuickFacts “Race and Hispanic Origin” Information were considered “White” to be consistent with the SRTR’s representation of “Hispanic/Latino” in their demographic records.

## Results

To measure the relationship between the representation of races/ethnicities across the waitlists for heart transplants, the heart transplants conducted, and the census (in the locations of the transplant centers), 3 Single-Factor ANOVA analyses were performed -- between the percent representation of all races/ethnicities on the waitlist, of those who received a transplant, and of the racial/ethnic representation of the cities in which these waitlists/transplants were conducted.

### Comparison Between Census Representation

A Single-Factor ANOVA analysis concluded that there was statistically significant variance between average racial/ethnic representation across all transplant centers (Fcalc = |175.0893697| > Fcrit = |2.385744612|) -- with statistical significance between “White” and “African-American” representation in the average census representation of all transplant centers (tcalc = |7.720743281| > tcrit = |1.969201329|), “White” and “Hispanic/Latino” representation (tcalc = |11.25765333| > tcrit = |1.969201329|), “White” and “Asian” (tcalc = |22.73824799| > tcrit = |1.969201329|), “White” and “Other” (tcalc = |25.86739437| > tcrit = |1.969201329|), “African-American” and “Hispanic/Latino” (tcalc = |2.846927984| > tcrit = |1.969201329|), “African-American” and “Asian” (tcalc = |11.38017033| > tcrit = |1.969201329|), African American and “Other” (tcalc = |13.56803315| > tcrit = |1.969201329|), “Hispanic/Latino” and “Asian” (tcalc = |8.868304545| > tcrit = |1.969201329|), “Hispanic/Latino” and “Other” (tcalc = |11.30798371| > tcrit = |1.969201329|), as well as between “Asian” and “Other” representation (tcalc = |4.265727343| > tcrit = |1.969201329|) using a Two-Sample Assuming Equal Variances tTest analyses.

**Figure 1:**
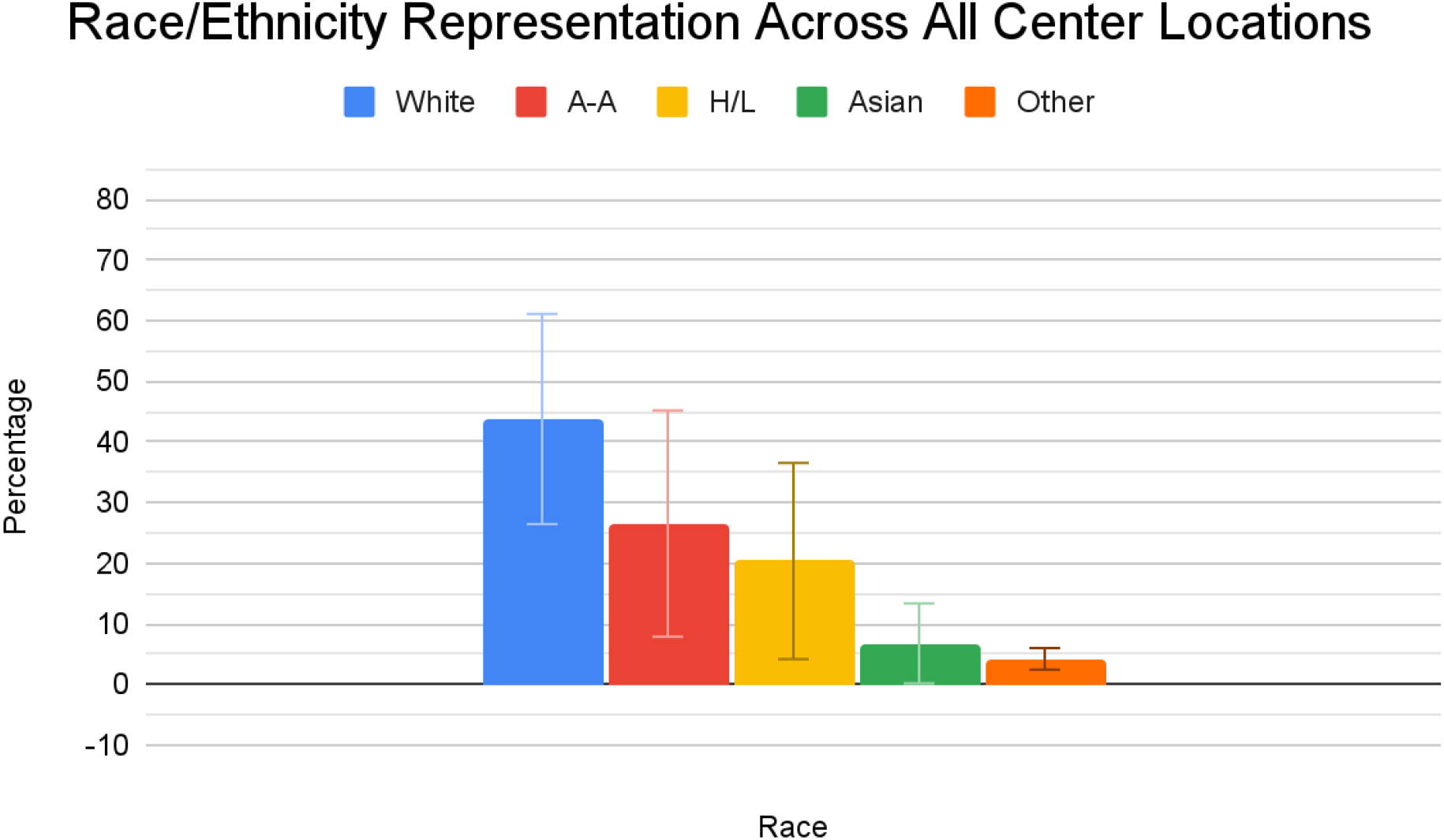
Comparison between all race/ethnicities’ average representation across all transplant centers’ locations where White = “White”, A-A = “African-American”, H/L = “Hispanic/Latino”, Asian = “Asian”, and Other = “Other” according to the United States Census Bureau QuickFacts (converted racial/ethnic representations for data compatibility with demographic data reportining by Scientific Registry of Transplant Recipients). Due to generalization across all centers, percentages may not add up to 100%.

### Comparison Between Waitlist Representation

Similarly, a Single-Factor ANOVA analysis concluded that there was a statistically significant variance between the representation of races/ethnicities on waitlists at all transplant centers that had waitlisted patients for heart transplants and that had passed trimming (Fcalc = |319.4644015| > Fcrit = |2.385744612|). Multiple Two-Sample Assuming Equal Variances tTest analyses were conducted and found that the differences between wait list representation between “White” and “African-American” (tcalc = |13.13408285| > tcrit = |1.969201329|), “White” and “Hispanic/Latino” (tcalc = |19.74497462| > tcrit = |1.969201329|), “White” and “Asian” (tcalc = |28.68676712| > tcrit = |1.969201329|), “White” and “Other” (tcalc = |30.70155265| > tcrit = |1.969201329|), “African-American” and “Hispanic/Latino” (tcalc = |6.160110656| > tcrit = |1.969201329|), “African-American” and “Asian” (tcalc = |12.733865718| > tcrit = |1.969201329|), “African-American” and “Other” (tcalc = |14.61603852| > tcrit = |1.969201329|), “Hispanic/Latino” and “Asian” (tcalc = |5.619981055| > tcrit = |1.969201329|), “Hispanic/Latino” and “Other” (tcalc = |7.59812063| > tcrit = |1.969201329|), as well as “Asian” and “Other” (tcalc = |4.455577492| > tcrit = |1.969201329|) were all statistically different from each “Other”.

**Figure 2:**
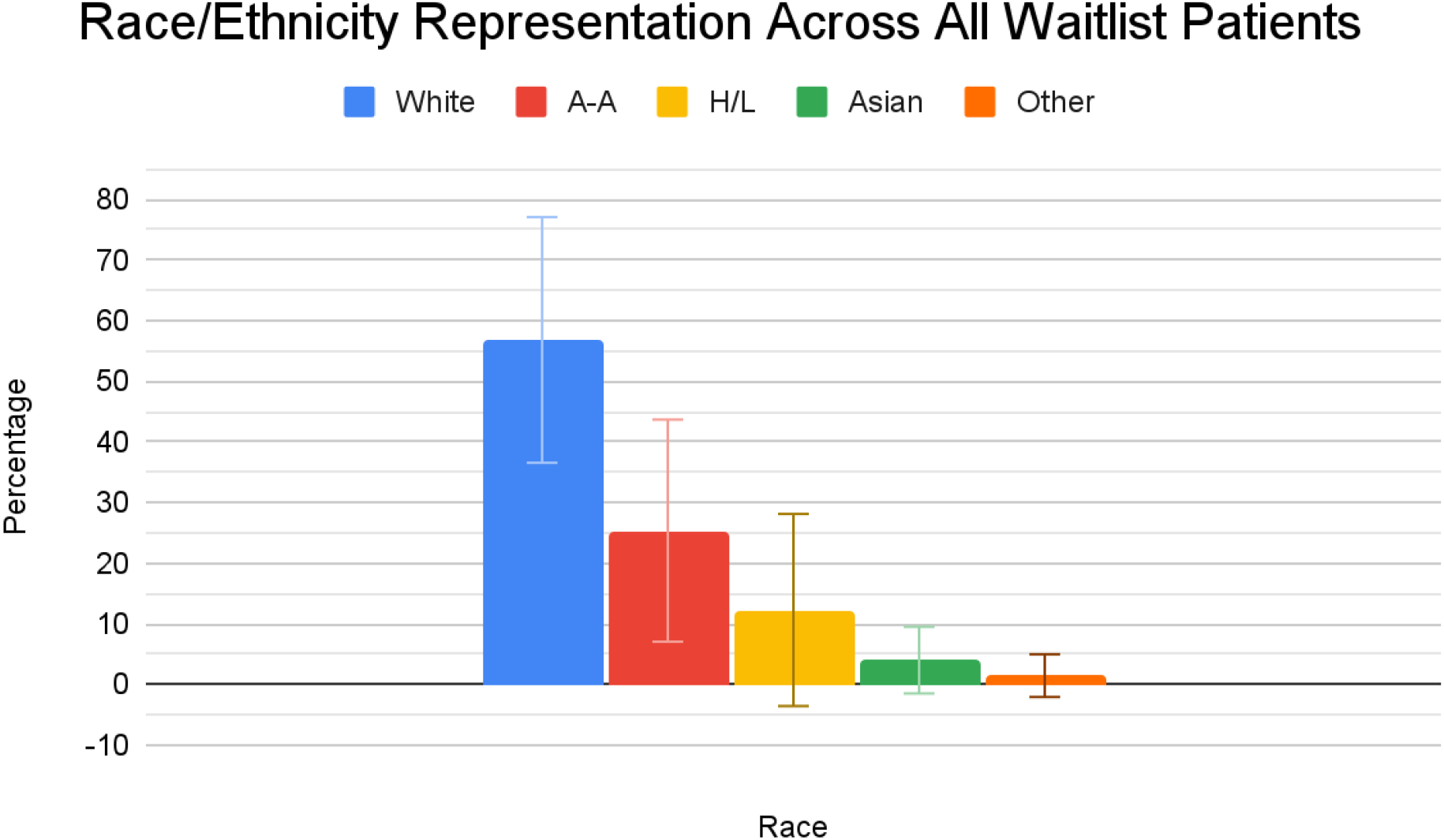
Comparison between all race/ethnicities’ average representation across all transplant centers’ waitlists for heart transplants where White = “White”, A-A = “African-American”, H/L = “Hispanic/Latino”, Asian = “Asian”, and Other = “Other”. Due to generalization across all centers, percentages may not add up to 100%.

### Comparison between Transplant Recipient Representation

An”Other” ANOVA analysis was performed on the racial/ethnic representation of all heart transplant recipients of transplant centers that passed trimming and found that there was a statistically significant amount of variance between races/ethnicities (Fcalc = |365.4409633| > Fcrit = |2.385744612|). Two-Sample Assuming Equal Variances tTest analyses concluded that the difference in racial/ethnic representation between “White” and “African-American” (tcalc = |15.65155112| > tcrit = |1.969201329|), “White” and “Hispanic/Latino” (tcalc = |21.33651592| > tcrit = |1.969201329|), “White” and “Asian” (tcalc = |29.00284189| > tcrit = |1.969201329|), “White” and “Other” (tcalc = |31.63965858| > tcrit = |1.969201329|), “African-American” and “Hispanic/Latino” (tcalc = |5.44149267| > tcrit = |1.969201329|), “African-American” and “Asian” (tcalc = |11.52652154| > tcrit = |1.969201329|), “African-American” and “Other” (tcalc = |14.0068482| > tcrit = |1.969201329|), “Hispanic/Latino” and “Asian” (tcalc = |5.45231139| > tcrit = |1.969201329|), “Hispanic/Latino” and “Other” (tcalc = |7.963016178| > tcrit = |1.969201329|), and “Asian” and “Other” (tcalc = |4.328597971| > tcrit = |1.969201329|) were significant as well.

**Figure 3:**
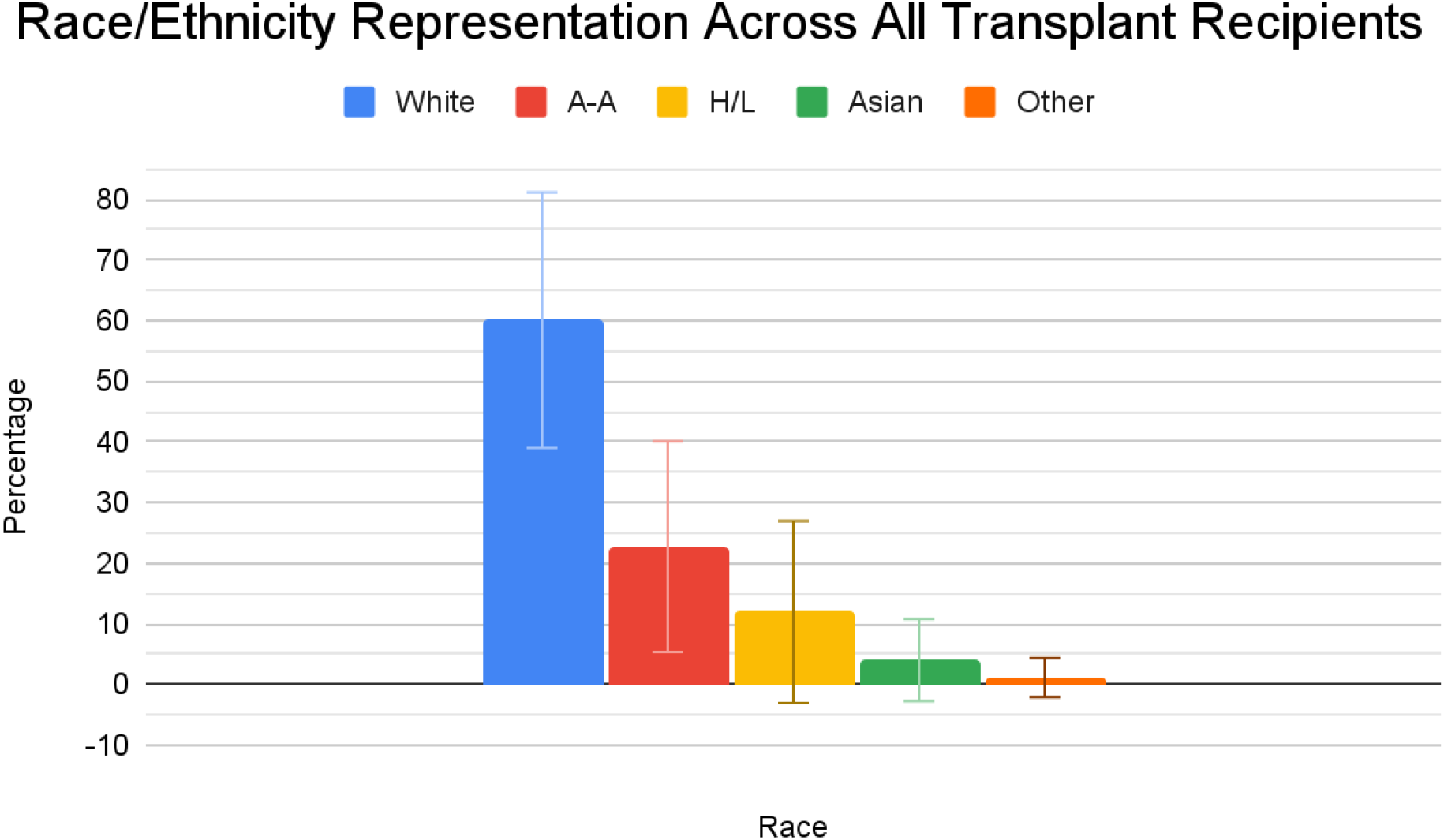
Comparison between all race/ethnicities’ average representation across the Census, Heart Transplant Waitlists, and Heart Transplant Recipients at all trimmed transplant centers, organized by race where WL = Waitlist Patient, T = Transplant Recipient, C = According to the United States Census Bureau QuickFacts (converted racial/ethnic representations for data compatibility with demographic data reportining by Scientific Registry of Transplant Recipients), White = “White”, A-A = “African-American”, H/L = “Hispanic/Latino”, Asian = “Asian”, and Other = “Other”. Due to generalization across all centers, percentages may not add up to 100%.

### Comparison between Transplant Recipient Representation

To see how various races/ethnicities varied across their representation in the census, heart transplant waitlists, and heart transplant recipients within their own race/ethnicity, 5 additional Single-Factor ANOVA analyses were performed.

#### “White” Representation

The variation between “White” representation across the census, heart transplant waitlists, and heart transplant recipients was found to be statistically significant (Fcalc = |25.42858764| > fcrit = |3.019042106|) with statistically significant differences between Census and Waitlist representation (tcalc = |5.586181963| > tcrit = |1.969201329|) and Census and Transplant representation (tcalc = |6.865205127| > tcrit = |1.969201329|). Representation between Waitlist and Transplant representation was statistically insignificant (tcalc = |-1.305828983| < tcrit = |1.969201329|).

#### “African-American” Representation

The variation between “African-American” representation across the census, heart transplant waitlists, and heart transplant recipients was not found to be statistically significant (Fcalc = |1.452066521| < fcrit = |3.019042106|).

#### “Hispanic/Latino” Representation

The variation between “Hispanic/Latino” representation across the census, heart transplant waitlists, and heart transplant recipients was found to be statistically significant (Fcalc = |12.13784941| > fcrit = |3.019042106|) with statistically significant differences between Census and Waitlist representation (tcalc = |-4.066739857| > tcrit = |1.969201329|) and Census and Transplant representation (tcalc = |-4.396773329| > tcrit = |1.969201329|). Representation between Waitlist and Transplant representation was statistically insignificant (tcalc = |0.222293716| < tcrit = |1.969201329|).

#### “Asian” Representation

The variation between “Asian” representation across the census, heart transplant waitlists, and heart transplant recipients was found to be statistically significant (Fcalc = |8.389453594| > fcrit = |3.019042106|) with statistically significant differences between Census and Waitlist representation (tcalc = |-3.675955957| > tcrit = |1.969201329|) and Census and Transplant representation (tcalc = |-3.362594318| > tcrit = |1.969201329|). Representation between Waitlist and Transplant representation was statistically insignificant (tcalc = |0.02314121875| < tcrit = |1.969201329|).

#### “Other” Representation

The variation between “Other” representation across the census, heart transplant waitlists, and heart transplant recipients was found to be statistically significant (Fcalc = |43.24602009| > fcrit = |3.019042106|) with statistically significant differences between Census and Waitlist representation (tcalc = |-8.005625736| > tcrit = |1.969201329|) and Census and Transplant representation (tcalc = |-9.541272129| > tcrit = |1.969201329|). Representation between Waitlist and Transplant representation was statistically insignificant (tcalc = |0.7440881075| < tcrit = |1.969201329|).

**Figure 3.**
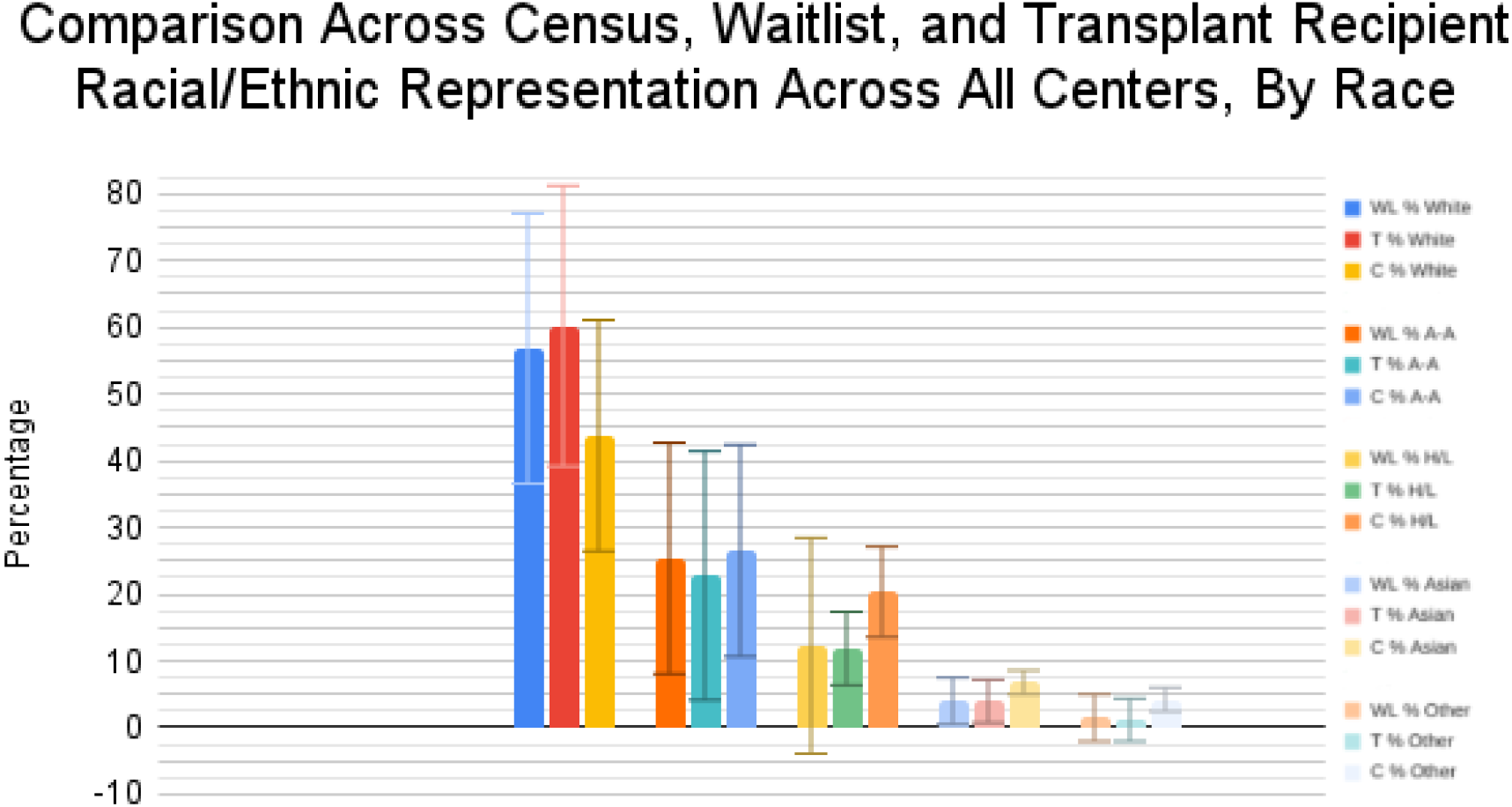
Comparison between all race/ethnicities’ average representation across the Census, Heart Transplant Waitlists, and Heart Transplant Recipients at all trimmed transplant centers, organized by race where WL = Waitlist Patient, T = Transplant Recipient, C = According to the United States Census Bureau QuickFacts (converted racial/ethnic representations for data compatibility with demographic data reportining by Scientific Registry of Transplant Recipients), White = “White”, A-A = “African-American”, H/L = “Hispanic/Latino”, Asian = “Asian”, and Other = “Other”. Due to generalization across all centers, percentages may not add up to 100%.

## Conclusions

With the exception of “African-American” individuals, all other ethnic/racial groups had significant differences in their representation in the average population in the areas in which the trimmed centers were located. “White” individuals, specifically, represented greater proportions of waitlist patients and transplant recipients compared to other races/ethnicities in relation to their demographic representation in the census, but those considered “Hispanic/Latino” and “Other” had the greatest degree of lop-sidedness to the representation of all racial/ethnic groups from waitlist/transplant recipient to census representation.

This points to many possible conclusions and/or confounding factors. For instance, “White” individuals may be more likely to seek out treatment than other races, be more likely to have access to care/the resources to seek out care (see Williams & Rucker, 2000), or they may be at higher risk for requiring heart transplants than other groups.

The reverse may be true for those in the “Asian”, “Hispanic/Latino”, and “Other” racial/ethnic groups; these individuals may be less likely to seek out care for various conditions (see Rojas-Guyler, King, & Montieth, 2008; Ben-Shlomo, Naqvi, & Baker, 2008; Warren-Jeanpiere, 2006), less likely to have access to healthcare services or the resources needed to seek it out (see Willging et al, 2018), more likely to adhere to medical regimens, and/or be at lower risk for requiring health transplantation in life, such as due to cultural diets and lifestyles (see Leu et al, 2019).

However, the lack of difference in representation amongst those in then “African-American” racial/ethnic category should not be interpreted as adequate ability to seek out and receive appropriate and equitable care. Black individuals are at higher risk for heart failure than nonblack individuals (Okin et al, 2011) and, given that heart failure often leads to requiring a heart transplant (Morris et al, 2016), the numbers for those who need to be placed on waiting lists and receiving transplants but do not turn up at centers may be higher. There are also other factors that may similarly affect other racial/ethnic groups such as a lack of trust in the healthcare system (see Jacobs et al, 2006; Cuevas & O’Brien, 2019), lack of access, awareness, or resources to pursue and attain care--amongst others.

It’s important to note that the data analyzed here is from transplant centers reports of waitlists and transplants from 07/01/2019 to 06/30/2020. The COVID-19 pandemic may have shifted numbers and demographic representation, especially in the latter half of the period.

Yet, this points to many potential avenues for future research and investigation: what influences can increase or decrease the likelihood of the onset of a condition that will require a heart transplant for the patient and how might we alter these influences to increase life expectancy and quality? How might we encourage populations who are at risk for requiring a heart transplant to seek out care at the appropriate time? How might individuals of different racial/ethnic groups view the value of seeking out care? How do perspectives between racial/ethnic groups surrounding the healthcare system, the use of transplants, or the value of extending life differ? How do other sub-cultural factors such as religion, sociocultural expectations, and values, morals, ideals, or ideas affect beliefs and attitudes around receiving transplants and/or care? How might specific cultural ideologies or thoughts such as the purpose of one’s life or the importance of continuing to live influence how willing someone is to accept treatment/waitlisting/transplantation?

Future research should also explore specific location-based trends in racial/ethnic representation in waitlists and transplants rather than a general look across all transplant centers. This was not possible in this study because of the lack of an adequate number of centers that qualified for the analysis in certain states--as 1-2 centers attempting to represent an entire state’s racial/ethnic representation would poorly represent efforts to provide equitable healthcare opportnities, especially in states with low population-density. By looking at specific location-based representations, more specific, population-based determinants of health for requiring a heart transplant/public health approaches needed to improve access to clinics/hospitals could be identified.

Additionally, due to the nature of racial/ethnic grouping, specific trends in subgroups that are more likely to have cardiovascular conditions that lead to heart transplants (e.g. South Asians) are unable to be seen under the general umbrella of an entire racial/ethnic group (e.g. “Asian”, which represents over 45 countries, with many having multiple unique sub-populations within them). Data reporting organizations should aim to diversify the number of racial/ethnic groupings to allow future researchers to better plot and understand sub-cultural changes and shifts in care, treatment, and transplantation over time.

## Data Availability

All data produced in the present study are available upon reasonable request to the authors.

## Acknowledgments

A special thanks to Dr. Kelsey Berry of the Harvard Medical School Center for Bioethics for the inspiration of this paper. Dr. Berry’s experiences and advice allowed me to understand the importance of understanding the demographic distributions of healthcare resources, especially in transplantations.

